# Fine-tuning Large Language Models in Behavioral Psychology for Scalable Physical Activity Coaching

**DOI:** 10.1101/2025.02.19.25322559

**Authors:** Sriya Mantena, Anders Johnson, Marily Oppezzo, Narayan Schuetz, Alexander Tolas, Ritu Doijad, C. Mikael Mattson, Allan Lawrie, Mariana Ramirez-Posada, Eleni Linos, Abby C. King, Fatima Rodriguez, Daniel Seung Kim, Euan A. Ashley

## Abstract

Personalized, smartphone-based coaching improves physical activity but relies on static, human-crafted messages. We introduce My Heart Counts (MHC)-Coach, a large language model fine-tuned on the Transtheoretical Model of Change. MHC-Coach generates messages tailored to an individual’s psychology (their “stage of change”), providing personalized support to foster long-term physical activity behavior change. To evaluate MHC-Coach’s efficacy, 632 participants compared human-expert and MHC-Coach text-based interventions encouraging physical activity. Among messages matched to an individual’s stage of change, 68.0% (N=430) preferred MHC-Coach-generated messages (*P <* 0.001). Blinded behavioral science experts (N=2) rated MHC-Coach messages higher than human-expert messages for perceived effectiveness (4.4 vs. 2.8) and Transtheoretical Model alignment (4.1 vs. 3.5) on a 5-point Likert scale. This work demonstrates how language models can operationalize behavioral science frameworks for personalized health coaching, promoting long-term physical activity and potentially reducing cardiovascular disease risk at scale.

## Introduction

Physical inactivity is one of the strongest risk factors for nearly all chronic diseases of aging.^1,2^ Despite the well-documented benefits of regular physical activity, a majority of adults in the United States remain sedentary.^3^ Mean daily steps is a commonly used proxy for physical activity, with even modest, short-term increases (+500 steps) associated with cardiovascular benefits.^4^ However, recent large-scale studies show that the average American achieves only 4,700 daily steps, well below the recommended threshold for health benefits.^4,5^ These findings emphasize the need for scalable interventions that effectively increase physical activity and support sustained engagement in the long term.

Personalized coaching has consistently demonstrated efficacy in increasing physical activity by addressing individuals’ unique needs, motivations, and barriers.^6–8^ Yet, sustaining long-term behavior change is a complex and dynamic process shaped by multiple interacting factors. Behavioral science offers structured, evidence-based frameworks for personalization to the individual by addressing the psychological, social, and environmental factors that influence behavior change.^9^ One well-established framework is the Transtheoretical Model (TTM),^10^ which focuses on the stages of change individuals undergo when adopting new healthy behaviors. The TTM categorizes individuals into five distinct stages of change: precontemplation, contemplation, preparation, action, and maintenance.^10^ The TTM has proven effective in guiding behavior change for chronic disease prevention, with tailored interventions at each stage demonstrating higher efficacy in increasing physical activity.^11^ Because the model’s interventions evolve with users as they progress through the stages of change (from precontemplation to maintenance), it supports both short- and long-term improvements in physical activity.^10^ Moreover, coaching strategies grounded in the TTM framework have fostered sustained physical activity across diverse populations.^12–14^ Despite its success in fostering behavior change, TTM-based coaching has largely relied on manual creation and delivery, limiting scalability. Digital health tools, such as smartphone apps and wearables, offer a promising avenue for expanding access to TTM-based interventions.

Digital health tools, particularly smartphone applications and wearable devices such as smartwatches, have been employed to encourage physical activity.^15,16^ Interventions using smartphone applications and fitness trackers have demonstrated a positive effect on physical activity, with a recent meta-analysis reporting an average increase of 1,850 steps per day when digital interventions include personalized feedback and notifications.^17^ While these findings highlight the potential of digital interventions, existing tools are currently limited in that they focus on short-term behavior change, with a lack of attention on sustained engagement and long-term impact. As a result, their effects on chronic disease prevention and treatment, such as cardiovascular disease, remain unknown. Moreover, while several digital trials are being conducted to assess the efficacy of TTM-based personalized coaching, they remain constrained by using preprogrammed, static messages designed by human experts and localized, center-based recruitment, limiting their scalability and adaptability.^18,19^

To address this issue of scale, recent digital health research has leveraged large language models (LLMs) to deliver health coaching. Generic LLMs can generate responses with high levels of empathy, readability, and helpfulness, often rivaling or exceeding those of human coaches.^20–23^ However, these systems are reactive, responding to user-initiated queries rather than using domain-specific reasoning in behavioral psychology to provide timely and motivation-sensitive interventions necessary for sustained behavior change. Meanwhile, Just-in-Time Adaptive Interventions (JITAIs) leverage real-time user data to deliver contextually relevant, timely support to enhance adherence and retention.^24^ However, these systems are limited to predefined decision rules and cannot generate behaviorally informed messaging. Hence, there remains a critical need to integrate behavioral science frameworks with LLMs to enable them to capture both the motivational language seen in chatbots and the adaptive, real-time responsiveness of JITAIs.

To explore this potential, we built upon the *My Heart Counts* (MHC) Cardiovascular Health Study, a large-scale digital health platform designed to promote physical activity. The *My Heart Counts Cardiovascular Health Study* was launched in 2015 as one of the first smartphone applications using Apple’s ResearchKit. On launch, it became the fastest-recruiting medical study of all time with >40,000 participants in the first two weeks.^25–30^ MHC is a smartphone-based observational study designed to gather frequent, remote data on fitness, activity, and sleep.^29^ With a later update, MHC leveraged the accessibility of smartphones and wearable technology to perform the first fully digital randomized trial, using a personalized, data-driven approach to encourage physical activity.^26^ In this randomized crossover trial using MHC, we recently demonstrated that personalized text-based coaching notifications, customized to each user’s baseline activity levels, significantly increased mean daily step count compared to all other digital notifications (e.g., a notification to reach 10,000 steps).^30^ However, this study was limited by scale, requiring human experts to craft each personalized coaching prompt a priori.^30^ Moreover, this study only considered short-term physical activity (over a 7-day intervention period), whereas the majority of cardiovascular benefits are seen with longer-term behavioral change.^30^

To address the challenges of scalability and the lack of long-term behavioral change in digital health research, we developed MHC-Coach, a fine-tuned LLM^31^ trained on TTM behavioral science principles^32^ and domain-specific knowledge in physical activity and cardiovascular health. MHC-Coach autonomously generates personalized, text-based coaching messages to increase physical activity, evolving with users’ changing needs, motivations, and readiness for behavior change. Unlike prior studies that relied on small evaluator groups (5 to 42 participants),^21–23,33,34^ we conduct a large-scale evaluation (N=632) with participants of the MHC study, comparing the efficacy of MHC-Coach messages vs. traditional human expert-designed messages. Additionally, behavioral science experts systematically rated both message types based on their motivational effectiveness and alignment with behavioral science principles. These analyses assess the potential of integrating behavioral science frameworks with AI-driven coaching to enhance personalization, improve motivation, and foster sustainable physical activity behavior change.

## Results

### 0.1 Fine-tuning MHC-Coach

MHC-Coach was developed by fine-tuning LLaMA 3-70B on a dataset of 3,268 human expert messages matched to a specific stage of change,^32^ a domain-specific corpus on health and exercise to provide background information on the importance of physical activity in preventing cardiovascular disease^1,35–41^, and detailed information about the TTM model and characteristics of individuals in each stage of change^10,42^ (see **Methods** and **Figure 1**). The combined fine-tuning dataset contained a total of 215,553 tokens.

**Figure 1.**
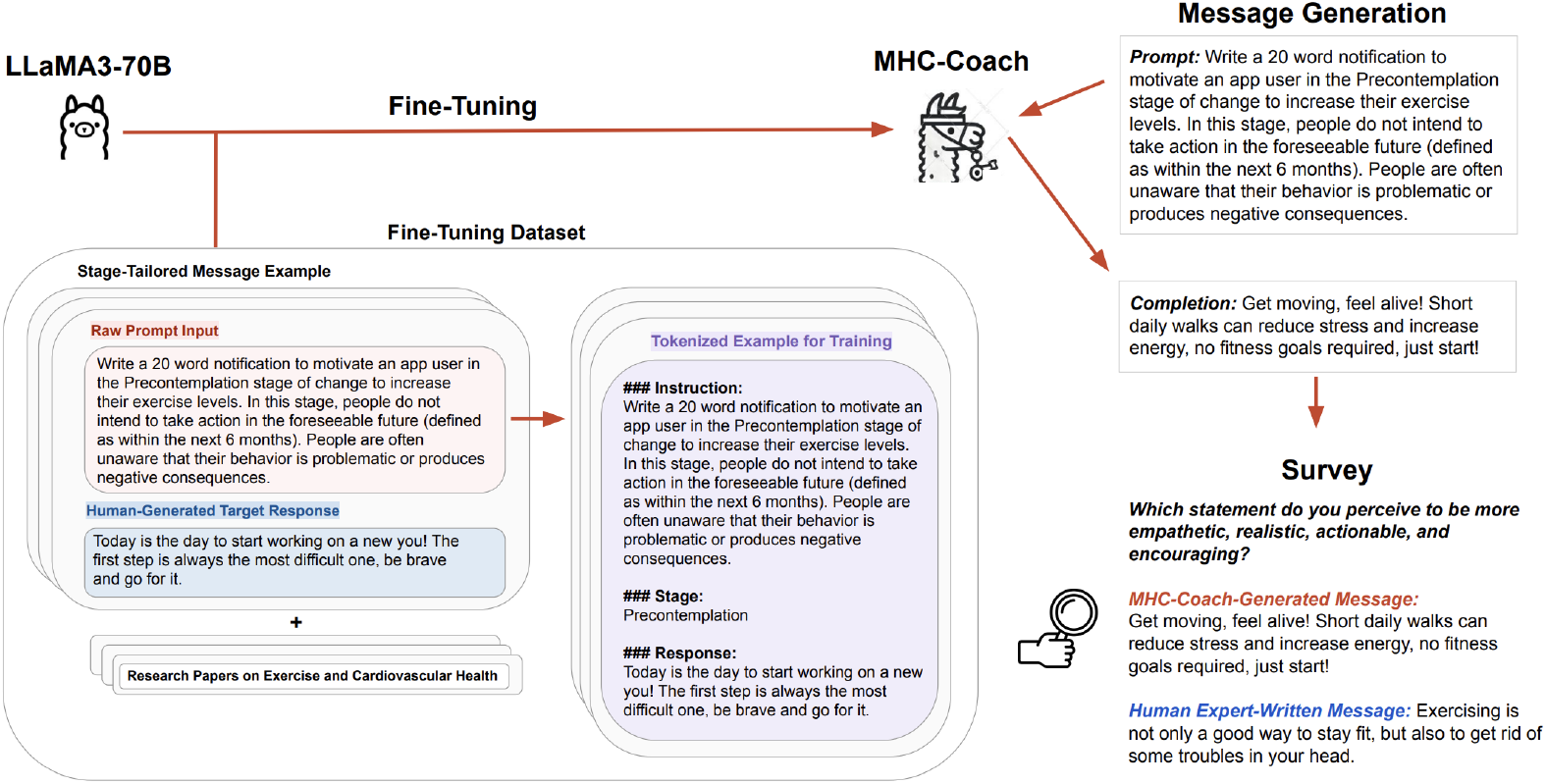
Schematic representation of the study design, including the fine-tuning process for LLaMA 3-70B, the generation of stage-specific motivational messages, and the evaluation through participant surveys.

To assess the effectiveness of this fine-tuning approach, we qualitatively compared MHC-Coach outputs to those of the base LLaMA 3-70B model, evaluating their integration of behavioral science principles and ability to generate stage-specific motivational messages (see **Figure 2**). For example, when asked to produce a motivation message for a user in the contemplation stage (i.e., those beginning to think about how to change their physical activity behavior), the base LLM generated:

**Figure 2.**
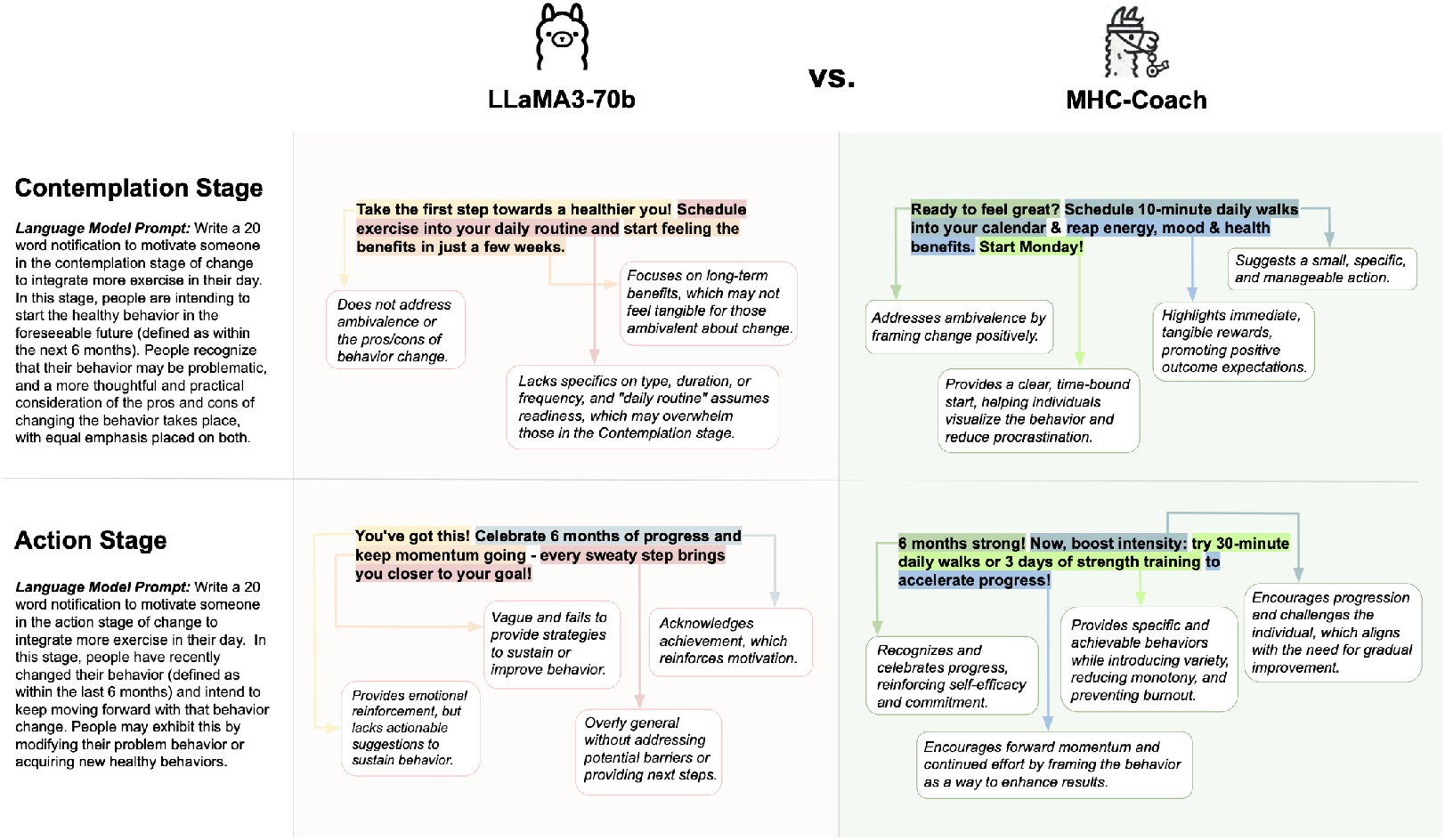
Qualitative comparison of Fine-Tuned LLM (MHC-Coach) and Base LLM (LLaMA3-70b) messages across the Contemplation and Action stages. Key differences include specificity in behavior recommendations and alignment with behavioral science principles, such as addressing ambivalence in the Contemplation stage and sustaining motivation through variety in the Action stage.

*“Take the first step towards a healthier you! Schedule exercise into your daily routine and start feeling the benefits in just a few weeks.”*

While this message encourages users to plan their exercise, it is overly general. It does not specify the type of exercise, the duration, or when to start, which makes it harder for users to visualize or act on the suggestion. In contrast, MHC-Coach generated the following message: *“Ready to feel great? Schedule 10-minute daily walks into your calendar & reap energy*, *mood, & health benefits. Start Monday!”*

This version demonstrates a significant improvement in actionable specificity by recommending “10-minute daily walks” and incorporating practical planning advice, such as integrating the activity “into your calendar.” The use of immediacy (*“Start Monday!”*) gives a clear starting point, while the explicit mention of benefits such as “energy, mood, & health benefits” creates a stronger connection between the action and its immediate outcomes. By combining actionable specificity, real-life planning, and outcome clarity, the fine-tuned message effectively integrates TTM behavioral science principles, making it more likely to resonate with and motivate users to take action. **Figure 2** illustrates these improvements with annotated examples from the contemplation and action stages of the TTM.

To further evaluate the effectiveness of MHC-Coach, we conducted two systematic assessments. The first was a survey-based preference study in which 632 prior MHC participants compared MHC-Coach messages to human coach-generated messages encouraging physical activity. In the second, behavioral science experts reviewed MHC-Coach and human expert messages and rated them on perceived effectiveness and TTM stage alignment.

### 0.2 Survey-based evaluation of MHC-Coach versus human expert text prompts

Survey invitations were sent via email to 62,616 users of the MHC smartphone app who had previously consented to re-contact. Surveys were conducted using the Qualtrics platform, and only participants who completed the entire survey were included in the analysis. Among the 1,004 initial survey respondents, 853 met the inclusion criteria of being at least 18 years old and English-speaking. Of these 853 eligible participants, 632 provided complete responses and were included in the final analysis (see **Figure 3**). The survey respondents included 79.4% males and 19.8% females, with the majority of participants (50.7%) aged 60+ years. The racial/ethnic distribution was predominantly White (80.8%), with smaller proportions identifying as Asian (6.1%), Hispanic/Latino (4.3%), and Black/African American (1.6%). Demographic characteristics of survey participants are summarized in **Table 1**.

**Table 1.** Participant Demographics: Summary of the study population’s demographic characteristics, including age, gender, ethnicity (with the option to select multiple ethnicities), and native language.

| Category | Subcategory | Count, % |
| --- | --- | --- |
| **Gender** | Male | 502, 79.4% |
|  | Female | 125, 19.8% |
|  | Non-Binary/Third Gender | 4, 0.6% |
|  | Prefer not to say | 1, 0.2% |
| **Age Group** | 10-19 | 2, 0.3% |
|  | 20-29 | 14, 2.2% |
|  | 30-39 | 63, 10.0% |
|  | 40-49 | 106, 16.8% |
|  | 50-59 | 127, 20.1% |
|  | 60+ | 320, 50.7% |
| **Ethnicity** | American Indian or Alaska Native | 14, 2.1% |
|  | Asian | 41, 6.1% |
|  | Black or African American | 11, 1.6% |
|  | Hispanic, Latino, or Spanish Origin | 29, 4.3% |
|  | White | 542, 80.8% |
|  | Other | 34, 5.1% |
| **Native Language** | English | 582, 92.1% |
|  | Spanish | 12, 1.9% |
|  | German | 5, 0.8% |
|  | Arabic | 6, 0.9% |
|  | Italian | 4, 0.6% |
|  | Other | 23, 3.6% |

**Figure 3.**
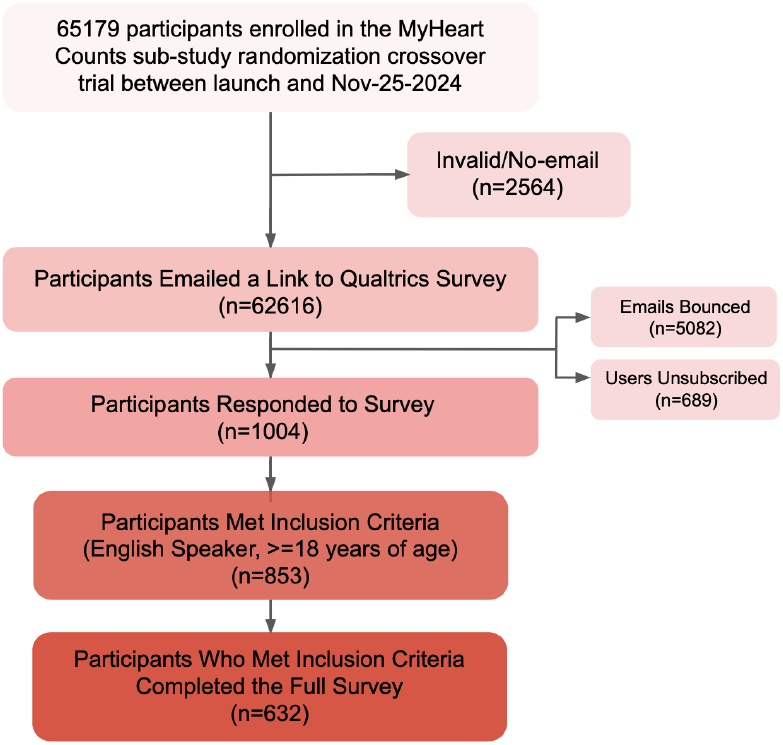
The recruitment process for the My Heart Counts study by comparing LLM-generated messages with expert-designed messages. Of 65,179 My Heart Counts participants who indicated openness to being contacted for future research, 62,616 were emailed the survey. A total of 1,004 participants responded, of which 853 met the inclusion criteria (English speakers aged *≥* 18 years), and 632 of the eligible participants completed the entire survey and were included in the final analysis.

To evaluate the efficacy of MHC-Coach in generating motivational messages, we surveyed participants with a series of forced-choice A/B questions. **Figure 4** illustrates the overall survey flow along with example questions. All participants received five forced-choice A/B questions comparing general motivational messages that were not tailored to a specific stage of change, with 85.4% (N=540) expressing a preference for the general MHC-Coach-generated message and 14.6% (N=92) preferring the human expert-designed message. Across the general motivational messages, there was a significant preference for the MHC-Coach-generated messages (Chi-squared *P <* 0.001, see **Table 2** and **Figure 5**).

**Table 2.** Preferences over coaching messages generated by LLMs versus expert-crafted messages, stratified by stages of change.

| TTM Stage of Change | N | Prefer LLM (N, %) | Prefer Expert (N, %) | Chi-Square P-Value |
| --- | --- | --- | --- | --- |
| Precontemplation | 37 | 27, 73.0% | 10, 27.0% | 0.005 |
| Contemplation | 89 | 64, 71.9% | 25, 28.1% | <0.001 |
| Preparation | 112 | 73, 65.2% | 39, 34.8% | 0.001 |
| Action | 76 | 61, 80.3% | 15, 19.7% | <0.001 |
| Maintenance | 318 | 205, 64.5% | 113, 35.5% | <0.001 |
| **Total** | **632** | **430, 68.0%** | **202, 32.0%** | **<0.001** |

| Message Type | N | Prefer General LLM Message (N, %) | Prefer General Coaching Message (N, %) | Chi-Square P-Value |
| --- | --- | --- | --- | --- |
| Generic Message | 632 | 540, 85.4% | 92, 14.6% | <0.001 |

**Figure 4.**
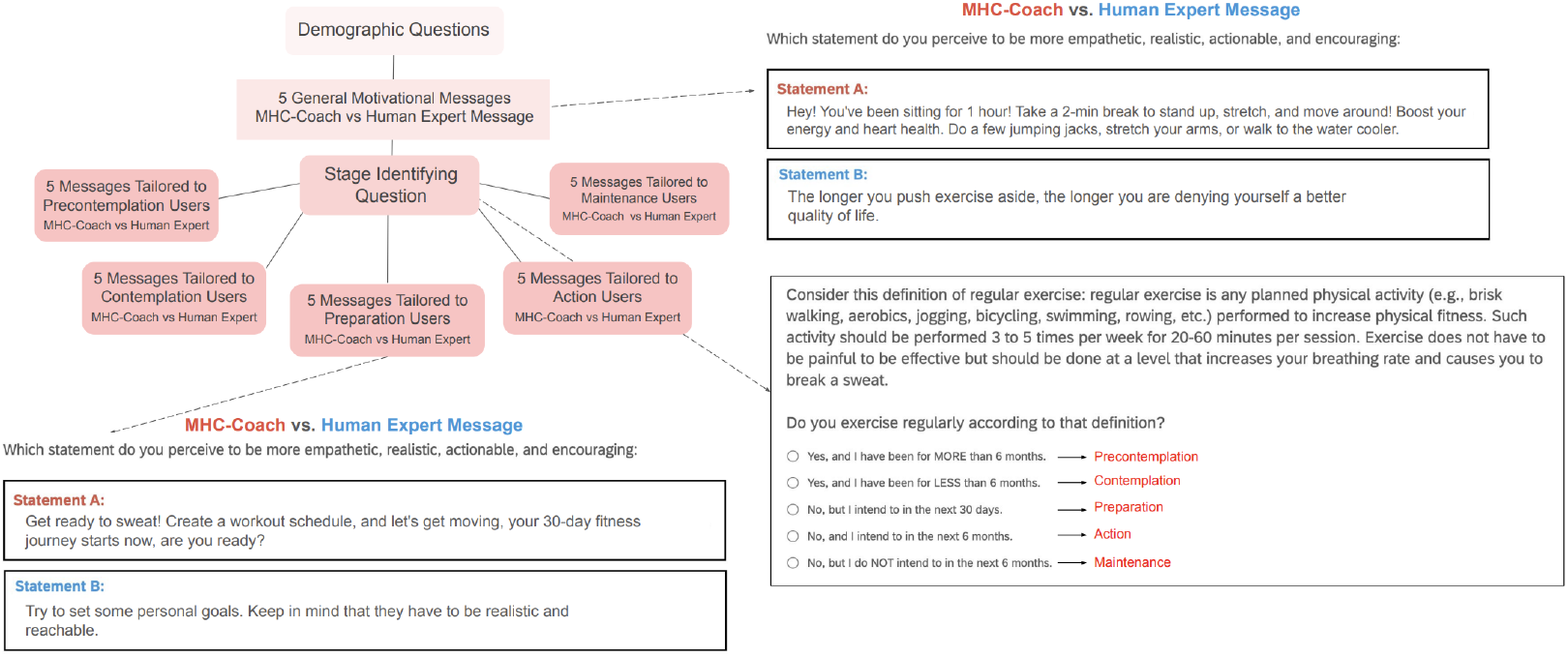
Overview of the survey design for comparing user preferences between LLM-generated and human expert-designed motivational messages. Users answered demographic questions and were asked to choose the prompt that they preferred on evaluation of 10 total message pairs, including 5 generally motivational messages and 5 pairs tailored to each user’s specific stage of change.

**Figure 5.**
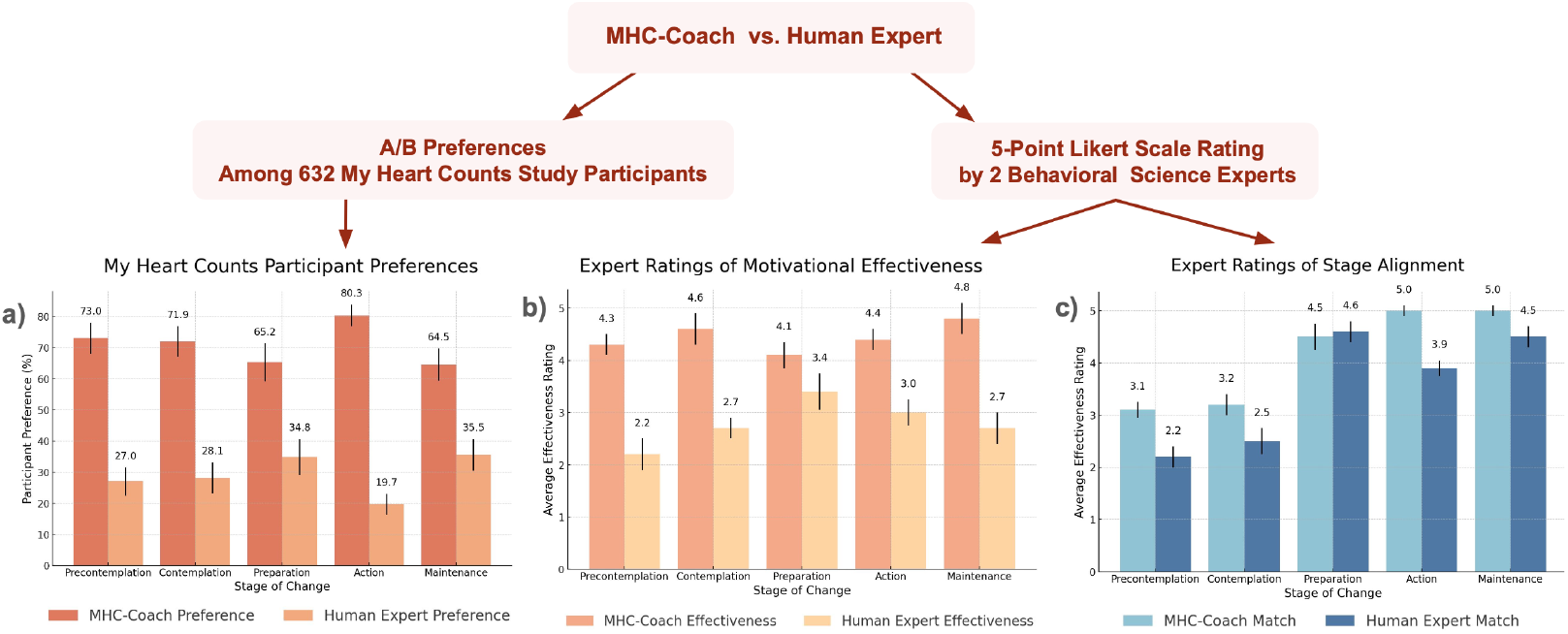
Comparisons between human expert-generated messages and fine-tuned LLM-generated messages across multiple evaluations. (a) Preferences of 632 surveyed users of the MyHeart Counts platform indicate a higher preference for LLM-generated messages across all stages of change. (b) Behavioral science experts rated the perceived effectiveness of messages on a 5-point Likert scale, with the fine-tuned LLM consistently outperforming human expert-generated messages. (c) Behavioral science experts evaluated how well each message aligned with the corresponding stage of change, with the fine-tuned LLM again achieving higher scores in most categories. Vertical lines for each bar represent the standard deviation.

Users were then stratified to a specific TTM stage of change and received three forced-choice A/B questions on which message they preferred: an MHC-Coach-generated or a human expert-created coaching message, matched to their individual stage of change. Across the stage-matched messages, 68.0% (N=430) of users showed a preference for the MHC-Coach-generated messages, and 32.0% (N=202) preferred the human expert messages (Chisquared *P <* 0.001, see **Table 2**). This preference was significant and consistent across all stages of change in a sensitivity analysis, with sample sizes ranging from *N* = 37 for the pre-contemplation stage to *N* = 318 for the maintenance stage. Detailed breakdowns of preferences across specific stages of change, along with associated statistical results, are presented in **Table 2** and **Figure 5**.

### 0.3 Behavioral science expert evaluation of MHC-Coach and human messages

Two behavioral science experts reviewed and rated the MHC-Coach-generated messages and human expert-written messages (see **Figure 5**). Across all stages of change, the average perceived effectiveness rating was 2.8 for the human-written messages and 4.4 for the MHC-Coach-generated messages on a 5-point Likert scale. Similarly, the average rating for how well the messages matched the requested TTM stage of change was 3.5 for the human-written messages and 4.1 for the MHC-Coach-generated messages. The most pronounced difference occurred in the pre-contemplation stage, where the human-written messages received an effectiveness rating of 2.2 and a match rating of 2.2, compared to the MHC-Coach-generated messages, which scored 4.3 for effectiveness and 3.1 for the stage match. A detailed breakdown of these ratings across all stages of change is presented in **Figure 5**.

## Discussion

Digital health offers the potential of a paradigm shift in scalability and individualization for physical activity coaching.^43^ Here, we developed and tested a tool, MHC-Coach, with substantial potential to address the gap in personalization via fine-tuning of a large language model to incorporate an evidence-based and validated psychological framework for behavior change. We show that MHC-Coach can provide autonomous, proactive, and personalized text-based coaching interventions, based on a user’s underlying behavioral psychology. Finally, we showed that MHC-Coach-generated messages were judged preferable by both behavioral science experts and participants in the *My Heart Counts* Cardiovascular Health Study, as compared to human expert-curated messages.

A key aspect of this work is demonstrating how fine-tuning LLMs with behavioral science principles enhances their ability to drive behavior change. Generic LLMs, despite their fluency and coherence, lack an inherent understanding of psychological motivation, resulting in responses that may be readable but not effective for long-term engagement. By embedding the TTM framework into an LLM’s latent space, the MHC-Coach model delivers structured, stage-specific coaching tailored to users’ behavioral readiness, surpassing both the base LLM and the human expert messages it was trained on. Moreover, with behavioral science as a backbone, future iterations of MHC-Coach can integrate existing personalization techniques based on activity habits, environmental context, and language preferences to further tailor interventions to individual needs.^24,44–46^ This could potentially bridge the gap and allow for ever-more-personalized and proactive LLM-generated health coaching at scale.

We further note that our study is unique amongst studies of LLM and health behavior for its scale and recruitment approach, reaching and engaging 632 MHC users seeking health interventions in the final analysis. This contrasts with the majority of preceding LLM-based health coaching studies, which relied on small, pre-selected groups of fewer than 42 annotators.^22,23^ We have collected user preference data and have made it openly available for further development of LLM-based health coaching leveraging the TTM framework (see **Methods**).

A few limitations of our study should be noted. First, our cohort reflects the user characteristics of the original *My Heart Counts* Cardiovascular Health Study and may not fully capture the diversity of the broader United States population.^25–30^ Moreover, MHC-Coach-generated messages were in English and hence restricted participation to English speakers. Research is ongoing to assess the efficacy of MHC-Coach-generated messages in non-English speakers, of which there are significant numbers within the United States. Finally, while MHC-Coach was fine-tuned on scientific health data to enhance accuracy, the possibility of hallucinating false or misleading information remains, a critical concern for health-related applications.^47,48^

In summary, MHC-Coach demonstrates the promise of integrating behavioral science frameworks with LLMs to deliver scalable, stage-specific health coaching. In contrast to generic LLMs, MHC-Coach goes beyond providing information about behavioral science and exercise; it operationalizes these principles in the language it generates, ensuring that coaching messages align with motivation-sensitive, stage-specific interventions. This study builds a foundation for future efforts, which include integrating MHC-Coach into the *My Heart Counts* smartphone application, expanding coaching to multilingual and culturally diverse settings, incorporating real-time activity data for automated stage classification, and evaluating long-term adherence and cardiovascular health outcomes through longitudinal studies.

## Methods

### 1. Message Generation

#### 1.1 Data

Our fine-tuning dataset consists of motivational messages based on the TTM framework, designed to promote exercise behavior change across the five stages of change, collected through a separate study.^32^ A total of 3,268 messages were included in this dataset, developed by 25 human experts, including fitness coaches, behavioral coaches, and researchers in health psychology, all of whom were native English speakers.^32^

In addition to these expert-designed messages, we incorporated content from manuscripts focused on the effects of regular exercise on cardiovascular health. The manuscripts^1,35–41^ contributed supplementary data that enriched the training corpus of MHC-Coach with evidencebased insights into the connection between physical activity and cardiovascular health outcomes, ensuring that the generated messages were scientifically grounded.

The combined fine-tuning dataset, consisting of human expert messages, content extracted from the research papers, and detailed information about the TTM model and characteristics of users in each phase,^10,42^ contained a total of 215,553 tokens. This dataset was utilized to finetune MHC-Coach, enabling it to generate contextually relevant and personalized messages for behavior change in physical activity (see **Figure 1**). By integrating expert-designed messages with research-based content, we aimed to ensure accurate health information aligned with clinical and behavioral science guidelines, while grounding the language in evidence-based psychology tailored to each TTM stage.

#### 1.2 Language Model

LLaMA 3-70B^31^ was used as the primary LLM architecture in this study. LLaMA 3-70B is a decoder-only transformer model with *>*70 billion parameters, optimized for diverse tasks such as reasoning, instruction following, and dialogue generation. For our experiments, we utilized LLaMA 3-70B with the following hyperparameters: temperature of 0.8 for response diversity, maximum token length of 800, and top-*p* (nucleus sampling) set to 0.9.

#### 1.3 Fine-tuning

We used an instruction-tuning^49^ variant to fine-tune LLaMA 3-70B on our dataset. Given the unstructured nature of the research papers and the need to align expert messages with the appropriate stage of change, we developed a custom template to guide the fine-tuning process. This custom template involved structuring the input data using a template designed to train the model to generate stage-specific motivational messages. Each example was designed with two key components: (1) a prompt section, describing the user’s stage of change and requesting a tailored motivational message, (2) a stage section specifying the user’s stage of change, and (3) a completion section, containing the expert-designed motivational message as the target response. To ensure clarity and consistency, a designated token was used to separate messages (see **Figure 1**). To provide additional context, unstructured text from manuscripts detailing the impact of regular exercise on cardiovascular health was appended to the training set,^1,35–41^ along with a comprehensive explanation of the TTM and the characteristics of individuals at each stage of change.^10,42^

We used Low-Rank Adaptation^50^ for fine-tuning, a technique that updates pre-trained models. This approach enables efficient fine-tuning by adding trainable low-rank matrices as additional parameters, leaving the original model’s weights unchanged. Fine-tuning was performed using a learning rate of 1 *×* 10^*−*5^, a batch size of 4, and 2 epochs. These parameters were determined through a grid-based hyperparameter search.

### 2. Expert Evaluation of Coaching Messages

We engaged two behavioral science experts to independently evaluate and rate both the fine-tuned MHC-Coach-generated messages and human-expert written messages. The evaluators were blinded as to whether a given message was MHC-Coach- or human-generated. They assessed each message on a 5-point Likert scale across two criteria: (1) the perceived effectiveness of the message in promoting increased physical activity and (2) the degree to which the message aligned with the recipient’s current stage of change.

### 3. Large-Scale User Study Design and Participant Recruitment

#### 3.1 Ethics

Ethical approval for the study was obtained from Stanford University’s Research Compliance Office (IRB-75836).

#### 3.2 Participant Recruitment and Eligibility Criteria

The *My Heart Counts* (MHC) smartphone application was first launched in March 2015 via the Apple App Store for United States, United Kingdom, and Hong Kong residents.^25–30^ Individuals aged 18 years or older who could read and understand English were eligible to participate in the *My Heart Counts* Cardiovascular Health Study.

Survey invitations were sent via email to MHC users who had indicated an openness to being contacted for future research. Survey responses were collected using the Qualtrics survey platform. Respondents who did not complete the full survey were excluded from the analysis, as complete responses were required for group-level analyses. Of the initial 1,004 respondents, 632 met the inclusion criteria, completed the entire survey, and were retained for the final analyses (see **Figure 3**).

#### 3.3 Survey Design

Throughout the survey, participants evaluated messages using an A/B forced-choice format, comparing pairs of messages—one human expert-designed and one generated by MHC-Coach—based on which they found more empathetic, realistic, actionable, and encouraging. The messages designed by human experts used for comparison were sourced from the same crowdsourcing study that provided the dataset for fine-tuning, involving 25 human specialists in fitness, behavior, and health psychology.^32^ Examples of both human expert-crafted and MHC-Coach-generated messages that users compared are provided in **Supplemental Table 1**. The first set of questions asked participants to choose their preferred text-based coaching prompt in an A/B forced-choice set of five questions featuring general motivational messages.

These messages were created by a human expert or generated by MHC-Coach with the sole objective of encouraging regular physical activity, independent of the stage of change.

Participants were then categorized into one of five stages of change based on their responses to an evidence-based question for TTM stage stratification assessing the duration and consistency of their regular exercise habits.^51^ Regular exercise was defined as planned physical activities—such as brisk walking, aerobics, jogging, bicycling, swimming, or rowing—performed 3 to 5 times per week for 20–60 minutes per session to improve physical fitness.^51^ After categorization, participants answered five questions comparing messages tailored to their specific stage of change. In this section, MHC-Coach-generated messages were crafted to correspond to the motivational and behavioral traits specific to each stage of change (see **Figure 1**).

#### 3.4 Statistical Analysis

The Chi-squared test was used to statistically evaluate differences in participants’ preferences for human expert-designed versus MHC-Coach-generated messages, under the null hypothesis that preferences for both types of messages were equal. Analyses were conducted across the five specific stages of change (pre-contemplation, contemplation, preparation, action, and maintenance) and for generic motivational messages. To account for the six comparisons performed (five stage-specific tests and one generic message test), a Bonferroni correction was applied to control the family-wise error rate, adjusting the significance threshold from *α* = 0.05 to *α*_adjusted_ = 0.0083 (0.05*/*6). Participants were classified as preferring MHC-Coach if they selected a majority of MHC-Coach-generated messages (3), and as preferring the human expert if they selected a majority of human expert-generated messages (3) in A/B forced-choice questions in analyses of the general motivational and TTM stage-specific question sets.

## Data Availability

The health coaching datasets generated and analyzed during the study are available on GitHub (https://github.com/SriyaM/MHC_LLM_Preference_Data). The dataset has been anonymized to ensure participant privacy, and no personally identifiable information is included. The repository includes code for data processing and customization to enable further analyses. Both the data and code are provided under the MIT License, allowing users to modify, reuse, and redistribute the materials in accordance with the license terms.

## Supplemental Materials

**Supplemental Table 1:** Examples of messages generated by the fine-tuned LLM Health Coach and human experts, tailored to different stages of behavior change. Expert messages were gathered from a crowdsourcing study involving 25 specialists in fitness, behavior, and health psychology.

| Stage of Change | Fine-Tuned LLM Health Coach Message | Human Expert Message |
| --- | --- | --- |
| Precontemplation | Get moving, feel alive! Short daily walks can reduce stress and increase energy, no fitness goals required, just start! | Try out something new today and surprise yourself with some physical activity. Who knows, maybe you will actually enjoy it? |
| Contemplation | Unleash your inner athlete! Exercise can help you discover new passions and interests. Try something new and have fun with it. | If others can do it, so can you. Try out a sport you haven't done before and figure out what you like the most. |
| Preparation | Get ready to sweat! Create a workout schedule, and let's get moving, your 30-day fitness journey starts now, are you ready? | Try to set some personal goals. Keep in mind that they have to be realistic and reachable. |
| Action | Get accountability and support! Share your fitness goals with a friend and schedule 30-minute walks together, 3 times a week. | It's often more fun to exercise together. Ask your friends to join you. |
| Maintenance | Remember why you started? Exercise daily to feel more energized, focused, and in control of your life. | If you continue now, feeling comfortable with your own body is going to be a steady aspect of your normal life. |
| Generic | Kick-start your day with a 10-min walk to reach 10,000 steps! Take a short break at work, walk to a coworker's desk, or do a few laps around your house. | The longer you push exercise aside, the longer you are denying yourself a better quality of life. |

